# Cardiorespiratory fitness and physical activity among children and adolescents with diabetes: A Systemic Review and Meta-analysis

**DOI:** 10.1101/2023.08.18.23294290

**Authors:** Hannah Steinman De Visser, Isaak Fast, Nicole Brunton, Edward Arevalo, Nicole Askin, Rasheda Rabbani, Ahmed M. Abou-Setta, Jonathan McGavock

## Abstract

**Background:** It is unclear if cardiorespiratory fitness (CRF) and physical activity (PA) are lower among youth with type 1 (T1D) and type 2 diabetes (T2D) compared to youth without diabetes. The objective of this study was to describe the magnitude, precision and constancy of the differences in CRF and PA between youth with and without diabetes.

**Methods:** We searched MEDLINE, Embase, CINAHL, and SPORTDiscus from 2000 to May 2022 for cross sectional studies that included measures of CRF and PA in children and adolescents with and without T1D or T2D. The main outcomes were objectively measured CRF obtained from a graded maximal exercise test and subjective or objective measures of PA. Three reviewers independently screened studies for eligibility, extracted data, and assessed studies for bias. Random effects meta-analysis model was used to estimate differences in main outcomes. The pooled effect estimate was measured as standardized mean differences (SMD) with its 95% confidence intervals (95%CI). This study was registered with PROSPERO (CRD42022329303) and followed the Preferred Reporting Items for Systematic Reviews and Meta-analyses (PRISMA) reporting guideline.

**Results:** Of 7857 unique citations retrieved, we included 9 studies (n = 755 participants) with measures of CRF and 9 studies (n = 1233 participants) with measures of PA for youth with T2D, as well as 23 studies (n = 2082 participants) and 36 studies (n = 12,196 participants) for youth with T1D. Random effects models revealed that directly measured CRF was lower in youth with T2D [SMD = - 1.06; 95% CI: -1.57 to -0.56, I^2^=84%, n=9 studies; n=781 youth] and in youth with T1D compared to controls [SMD = -0.39; 95% CI: -0.70 to -0.09, I^2^=89%, n=22 studies; n=2082 youth]. Random effects models revealed that daily PA was marginally lower in youth with T1D [SMD= -0.29, 95% CI −0.46 to -0.11, I^2^=89%, n=32 studies; n = 12196 youth] but not different among youth with T2D compared to controls [SMD= -0.56, 95% CI −1.28 to +0.16, I^2^=91%, n=9 studies; n=1233 youth]. When analyses were restricted to studies with objective measures, PA was significantly lower in youth with T2D [SMD -0.71, 95% CI −1.36 to -0.05; I^2^=23%, n=3 studies; n=332 youth] and T1D [SMD -0.67, 95% CI –1.17 to -0.17; I^2^=93%; n=12 studies; n=1357 youth] compared to controls.

**Conclusions:** Children and adolescents living with T1D and T2D display lower CRF and objectively measured PA compared to controls without diabetes. Deficits in CRF, but not PA are larger and more consistent in youth with T2D, compared to youth with T1D. These data may contribute to the increased risk for cardiovascular disease-related morbidity observed in adolescents with diabetes, particularly those with T2D.

**Funding:** This study was funded by the Canadian Institutes of Health Research

## INTRODUCTION

Cardiorespiratory fitness^1,2^ and daily physical activity^3,4^ are important modifiable determinants of pediatric and adult cardiometabolic disease, including mortality^5^. Adolescents with high cardiorespiratory fitness^6,7^ and those that engage in regular daily physical activity^3,4^ display lower blood pressure, lower central and overall obesity and are less likely to develop cardiovascular disease-related morbidity in adulthood. Accordingly, engaging in regular physical activity^8,9^, and by extension, increasing cardiorespiratory fitness^10^ are consistent recommendations for optimal growth and health for children and adolescents, including those living with diabetes^11–15^.

Current clinical practice and expert guidelines argue that children and adolescents (referred to herein as youth) with type 1 and type 2 diabetes are less active and have lower cardiorespiratory fitness than their peers without diabetes^11–15^. Despite these claims, the evidence is mixed. Some studies have documented large deficits in physical activity and cardiorespiratory fitness in youth with type 2 diabetes^16^ while others suggest differences are less profound^17^. Among youth with type 1 diabetes, some studies found lower levels of physical activity and cardiorespiratory fitness^18^, while many suggest no difference^19^ and in some cases, higher levels of physical activity^20^. In the absence of a systematic review and meta-analysis of observational studies, the consistency, magnitude and precision of these differences remain unclear. More accurate estimates of the differences in physical activity and cardiorespiratory fitness among youth living with chronic diseases is a priority among researchers in this area^21^ and the magnitude of these differences could help tailor behavioural interventions to improve health outcomes for youth living with diabetes.

To address these gaps in pediatric diabetes, we conducted a systematic review and meta-analysis to answer the following questions: Are cardiorespiratory fitness and physical activity lower among youth with diabetes compared to youth without diabetes? What is the overall quality of studies that describe differences in cardiorespiratory fitness and physical activity among youth with diabetes and controls without diabetes?

## METHODS

### Data sources, search strategy, and eligibility criteria

This review was conducted according to the Methodological Expectations of Cochrane Reviews and reported according to the PRISMA guidelines^22^. We searched MEDLINE (Ovid), Embase (Ovid), CINAHL (EBSCOhost), and SPORTDiscus (EBSCOhost). Controlled vocabulary and free text terms were utilized. A search for cross sectional and cohort studies was conducted from January 1, 2000 to May 1, 2022 (Supplementary eTables 1). The systematic review followed a priori eligibility criteria and the protocol was registered on PROSPERO (CRD:42022329303). Institutional ethics approval was not required for this systematic review and meta-analysis as individual-level data were not used for this analysis.

### Study inclusion criteria

We included cross sectional and cohort studies that compared measures of cardiorespiratory fitness or physical activity between youth with type 1 diabetes or type 2 diabetes and controls without diabetes. The inclusion criteria for data extraction and meta-analyses were: studies of youth aged 0 to 18yrs of age, cases of youth with type 1 diabetes or type 2 diabetes, a comparison group without diabetes, published estimates of differences and variance for physical activity or cardiorespiratory fitness between youth with diabetes and controls without diabetes. We excluded experimental studies, studies that included data from adults (over the age of 18), studies that compared youth with type 1 and type 2 diabetes directly without a comparison group without diabetes, and studies published in languages other than English.

### Classification of diabetes-types

We applied definitions established by Diabetes Canada^23^ and the American Diabetes Association^24^ to define type 2 diabetes and type 1 diabetes in youth.

### Study selection

Abstracts and titles of relevant citations were independently screened by three reviewers (HSD, EA, IF) to determine eligibility. Prior to the full screen, a preliminary screen of 30 titles and abstracts was conducted to determine a preliminary agreement rating. As the preliminary agreement rate was adequate (28/30 abstracts were in agreement), the same reviewers independently screened all remaining titles and abstracts. Two reviewers (HSD, IF) independently assessed the eligibility of full-text articles of citations using a standardized pre-piloted form outlining the inclusion and exclusion criteria. Disagreements were resolved by consensus or with the involvement of a third reviewer (JMc).

### Data extraction

Data were extracted independently by two reviewers, with disagreements resolved by consensus or with the involvement of a third reviewer. Two authors extracted data for study population demographics (age, sex, weight status, ethnicity, duration of diabetes, ethnicity, socio-economic status) and outcome measures. We also created binary variables for the methods used to collect physical activity (objective vs subjective) and cardiorespiratory fitness (field vs lab) and within the measure of cardiorespiratory fitness whether the tests were maximal or sub-maximal. Objective methods included pedometer or accelerometer-based quantification of daily physical activity, while subjective methods included a range of self-reported tools. Lab-based methods to quantify cardiorespiratory fitness included graded maximal tests to exhaustion and sub-maximal tests with measured rates of oxygen consumption. Field tests of cardiorespiratory fitness included graded maximal tests to exhaustion (i.e. shuttle run tests) that reported predicted peak rates of oxygen consumption or time to exhaustion.

### Outcome and subgroup analyses

Cardiorespiratory fitness was reported as the maximal rate of oxygen consumption relative to body weight (mL/kg/min) or fat free mass mass (ml/kg-FFM/min) or time to exhaustion at the end of a graded maximal exercise test. Physical activity was reported as minutes of physical activity per day, minutes of moderate to vigorous physical activity per day, METS, MET-mins and MET-hours per week. As there was significant heterogeneity for the methods and reporting of both cardiorespiratory fitness and physical activity, we used the J correction factor for an unbiased estimate of the standard mean difference (SMD) between youth with and without diabetes^25^.

### Risk of bias assessment

We evaluated the internal validity of included studies using a modified version of a risk of bias tool used in a previous meta-analysis of cross sectional studies of physical activity for youth living with chronic diseases^19^ (Appendix eTable 1). This risk of bias tool consists of several domains specific to measurements of physical activity: type of measurement (objective vs subjective), time of year for data collection, data collection and reduction of physical activity data, and criteria used to define a valid file for physical activity data. We also included several domains specific to the design of cross-sectional studies including a description of the sampling recruitment for cases and controls, eligibility criteria for controls, identifying and matching for potential sources of confounding, sex disaggregated analyses, and reporting of and matching for ethnicity. For studies of cardiorespiratory fitness, we also included a domain for reporting criteria for determining if a maximal effort was achieved and a score for the overall risk of bias. Each domain was judged as yes or no and given a score of 1 or 0 for the presence or absence of each potential source of bias. The overall risk of bias score was based on the responses to individual domains.

### Statistical analyses

The primary outcomes for the meta-analyses were standardized mean differences in physical activity and cardiorespiratory fitness. When at least 2 studies of similar populations, methods, and outcomes were available, inverse-variance weighted random-effects meta-analyses were performed to calculate standardized mean differences with 95% confidence intervals (95% CI) between youth with and without diabetes. We initially calculated standardized mean differences with 95% CI for all studies that met inclusion criteria. Absolute mean differences with 95% CI were calculated for differences in the main outcomes for sub-group analyses of studies that used identical tools to quantify outcome measures between youth with and without diabetes. The range of differences and the inconsistency index (I^2^) were both used to quantify heterogeneity among studies^25,26^. Subgroup analyses were performed to determine if differences in outcomes were influenced by: (A) the methods used to assess the main outcomes of interest (direct vs indirect measures for cardiorespiratory fitness and subjective vs objective measures for physical activity), and (B) the type of control population studied (healthy weight vs weight matched). We also performed sensitivity analyses by removing extreme outliers. Funnel plots were used to investigate publication bias with the Egger test and visual inspection to assess plot asymmetry for all four meta-analyses. The meta-analyses were conducted using the general meta and metafor package^27^ in RStudio version 2022.07.2+576, R version 4.2.2 (R Project for Statistical Computing).

## RESULTS

After systemic searches of each database, 7857 unique individual citations were included in a preliminary screen. At the end of the title and abstract screening, 7400 papers were excluded, 52 were included, and 405 were in conflict across the three reviewers. There was a 94.8% agreement rate between the reviewers for the title and abstract screen. The 405 papers with conflicting scores were resolved by the 3 screeners and one investigator (JMc) to determine consensus for eligibility for full paper screening and data extraction (see eTables 2 and 3 for references). After all the conflicts were resolved, 130 papers were included for the full paper screen and 55 studies were included for data extraction^1,5,6,16–18,20,28–77^ (eFigure 1 appendix). Details for each individual study are presented in eTables 2 through 5 of the appendix.

For studies of pediatric type 2 diabetes, most studies of physical activity (8/9) and cardiorespiratory fitness (8/9) were performed exclusively with adolescents. Among studies of cardiorespiratory fitness among youth with type 2 diabetes, the proportion of female participants was 56.19% for cases and 59.13% for controls, the median age was 15.3 years for cases (range: 12.9, 16.4 years) and 15.1 years for controls (range: 12.5, 16.2 years), and all participants were obese. Only 6 of 10 studies included information about the ethnicity of their participants (eTable 6 of the appendix).

For studies of physical activity among youth with type 2 diabetes, the proportion of females was 68.1% for cases and 58.7% for controls. The median age was 15.2 years (range: 12.9, 16.4) for cases and 14.8 years for controls (range: 12.5, 16), and all participants were obese. For the studies of pediatric type 2 diabetes that reported physical activity, 7 of the 9 studies included information about the ethnicity of their participants (eTable 6 of the appendix).

For studies of youth with type 1 diabetes, most studies of cardiorespiratory fitness (n = 17/23; 73.9%) and physical activity (n=24/36; 66.7%) were performed in adolescents. The proportion of female participants was 45.7% in cases and 49.7% in controls for studies of cardiorespiratory fitness and 51.6% in cases and 50.7% in controls for studies the reported physical activity. The median age was 14.1 years (range: 10.5 to 16.2 years) for cases and 13.6 years (range: 10.1 to 16.0 years) for controls in studies of cardiorespiratory fitness and 14.0 years (range: 7.4 to 16.0 years) for cases and 13.6 years (range: 7.3 to 16.3 years) for controls for studies of physical activity. The distribution of ethnicity for the studies that reported it is provided in eTable 7 of the appendix.

Among the 55 studies included in data extraction, 9 studies (n = 755 participants; 286 cases) met the eligibility criteria for the outcome of cardiorespiratory fitness for youth with type 2 diabetes and 22 studies (38 unique comparisons; n = 2082 participants; 1018 cases) met the eligibility criteria for the outcome of cardiorespiratory fitness for youth with type 1 diabetes. Mean VO_2_peak values from lab-based studies was 20.7 ± 3.5 ml/kg/min for youth with type 2 diabetes and 36.9 ± 5.3 ml/kg/min for youth with type 1 diabetes (Figure 1). The random-effects meta-analysis revealed that youth with type 2 diabetes displayed significantly lower cardiorespiratory fitness relative to controls without diabetes [SMD = -1.06; 95%CI: -1.57 to -0.56, I^2^=84%, n=9 studies; n=755 youth] (Figure 2A). When data were restricted to studies with lab-based measures of VO_2_peak, youth with type 2 diabetes displayed significantly lower cardiorespiratory fitness compared to controls without diabetes [MD= -8.76 ml/kg/min, 95% CI: -12.7 to -4.9; I^2^ = 82%; 8 studies; n= 575 participants].

**Figure 1.**
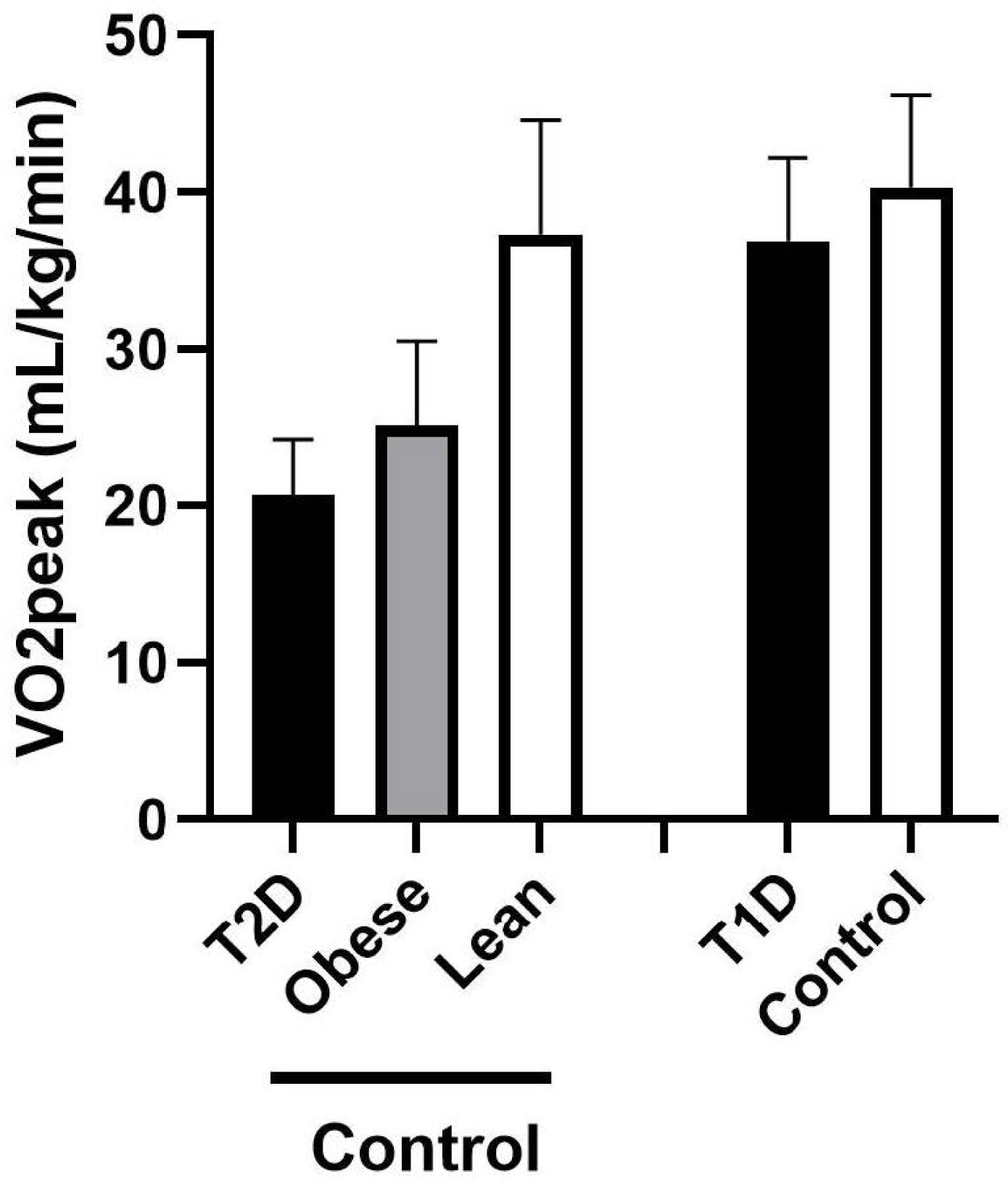
Differences in VO_2_peak between youth without diabetes compared to youth with type 1 and type 2 diabetes.

**Figure 2.**
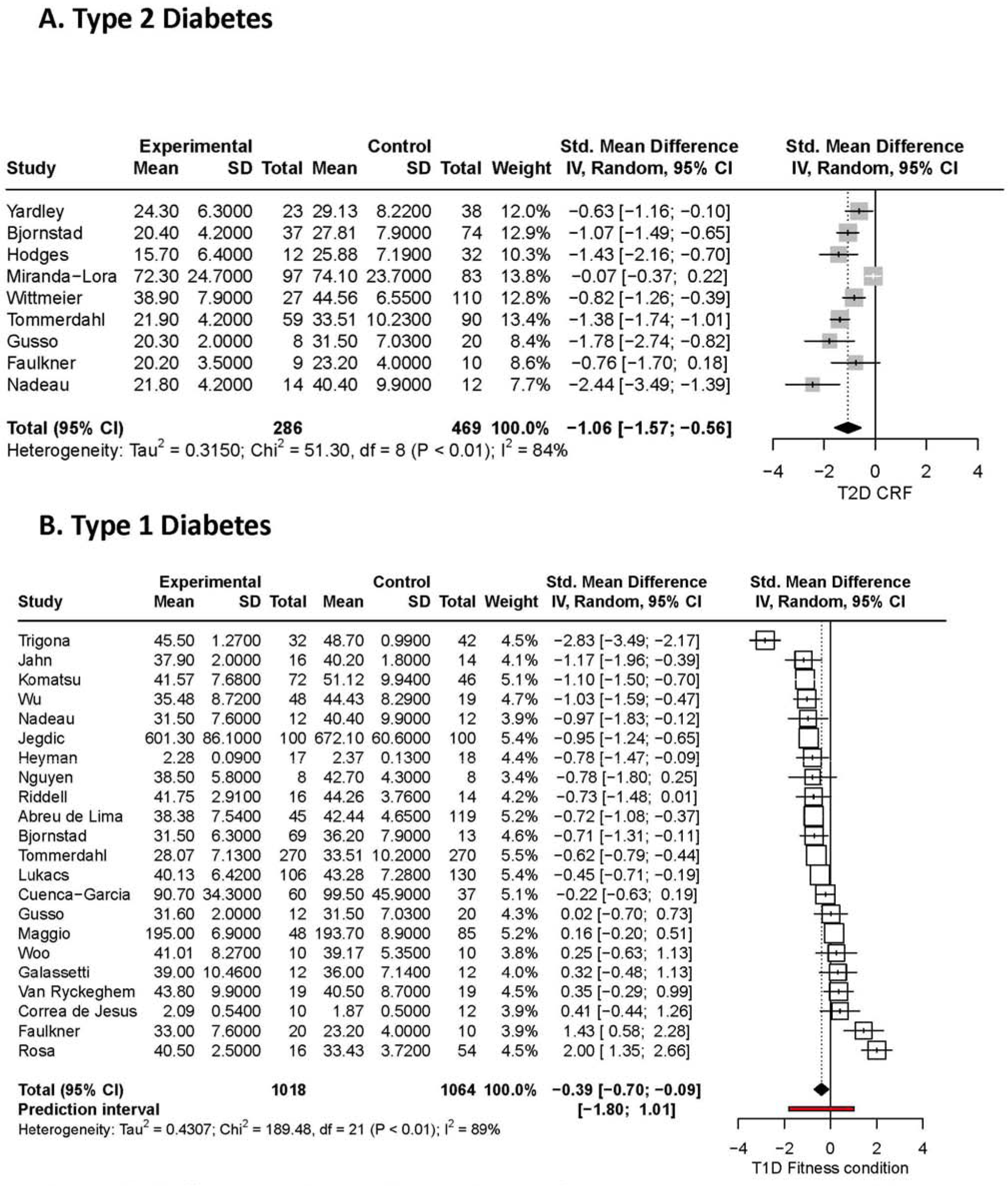
Differences in cardiorespiratory fitness between youth with diabetes and controls.

The random-effect meta-analysis revealed cardiorespiratory fitness was also lower in youth with type 1 diabetes compared to controls without diabetes [SMD = -0.39; 95% CI: -0.70 to -0.09, I^2^=89%; n= 23 studies; n=2082 participants; 1018 cases] (Figure 2B). Restricting the analysis to lab-based measures of cardiorespiratory fitness did not influence the estimate of the difference between youth with type 1 diabetes and controls without diabetes [MD: -3.9 ml/kg/min; 95% CI: -6.3 to -1.6 ml/kg/min; I^2^ = 68%; n=13 studies; n = 917 participants]. Subgroup analyses of studies of youth with type 2 diabetes revealed that the differences in cardiorespiratory fitness were lower when controls without diabetes were obese [SMD = -0.75; 95% CI:-1.15 to -0.35; I^2^ = 77%; n=8 studies; n= 593], compared to studies with controls that were healthy weight [SMD = -2.10 ; 95% CI: -2.74 to -1.47; I^2^ = 77%; n = 7 studies; n= 328]. Subgroup analyses restricted to health weight youth with type 1 diabetes, revealed that cardiorespiratory fitness was lower compared to healthy weight controls without diabetes [SMD = -0.75; 95% CI: -1.09 to -0.41; I^2^ = 86%; n = 15 studies; n= 1295 participants]. Only 2 studies were performed in youth with type 1 diabetes that were obese and no differences in cardiorespiratory fitness were observed relative to controls without diabetes.

Of the 55 studies that met eligibility for data extraction, 9 studies (10 unique comparisons; n = 1233 participants) met eligibility for the outcome of physical activity for youth with type 2 diabetes and 36 studies (n = 12,196 participants) met eligibility for the outcome of physical activity for youth with type 1 diabetes. Random effects models revealed that daily physical activity was not different among youth with type 2 diabetes [SMD= -0.56, 95% CI –1.28 to +0.16, I^2^=91%] (Figure 3A) studies compared to controls without diabetes, but was marginally lower among youth with type 1 diabetes [SMD= -0.29, 95% CI –0.46 to -0.11; I^2^ = 89%] (Figure 3B) compared to controls without diabetes. Analyses restricted to studies with objective measures found that physical activity was lower in youth with type 2 diabetes [SMD -0.71, 95% CI –1.36 to -0.05; I^2^ = 23%; n=3 studies; n=332] and type 1 diabetes [SMD -0.67, 95% CI –1.17 to -0.17; I^2^ = 93%; n=12 studies; n=1357] relative to controls without diabetes (eFigures 2 and 3).

**Figure 3.**
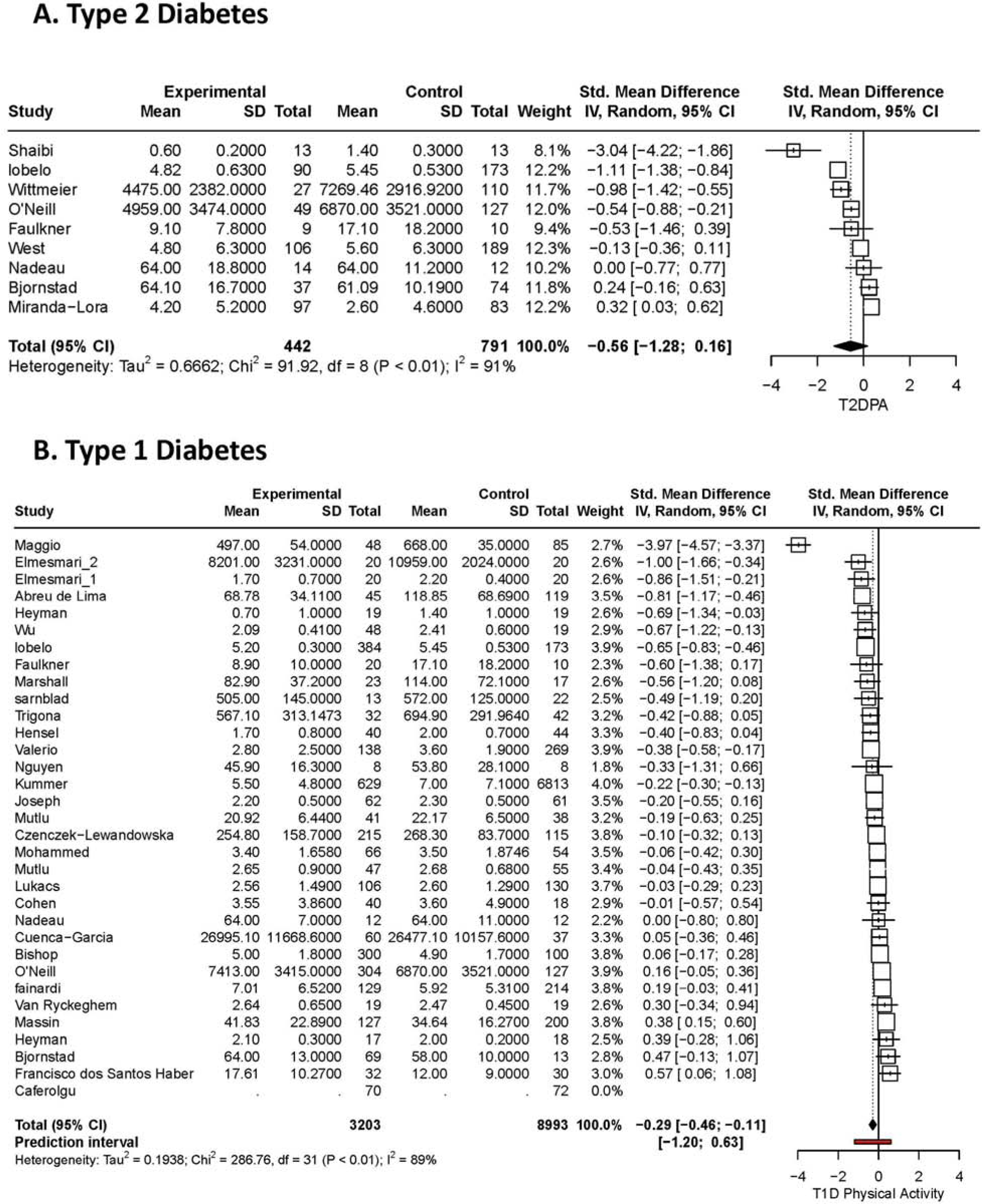
Differences in daily physical activity between youth with diabetes and controls.

The distribution of bias for studies of cardiorespiratory fitness and physical activity among youth with type 2 diabetes and type 1 diabetes are summarized in Table 2. For all studies combined, 80-89% of studies provided information on the recruitment process for the comparison population. Only 22-36% of studies described the timing of the measurements of physical activity, 20-39% of studies described methods to address potential sources of bias or strategies to address confounding and 0 to 22% presented sex disaggregated data. For studies of youth with type 2 diabetes, the majority (n=8/9) relied on lab-based measures of cardiorespiratory fitness. For studies of physical activity among youth with type 2 diabetes, only 3/9 studies used objective measures and none of the studies provided definitions for a valid assessment of physical activity or described the season during which data were collected. For studies of cardiorespiratory fitness among youth with type 1 diabetes, the majority relied on laboratory-based maximal tests to exhaustion (21/23) but few reported criteria for achieving maximal exertion. For studies of physical activity among youth with type 1 diabetes, 13/36 used objective measures and 61% provided definitions for a valid assessment of physical activity. Funnel plots revealed no publication bias for the four meta-analyses presented above (eFigures 4 through 7).

**Table 1.**
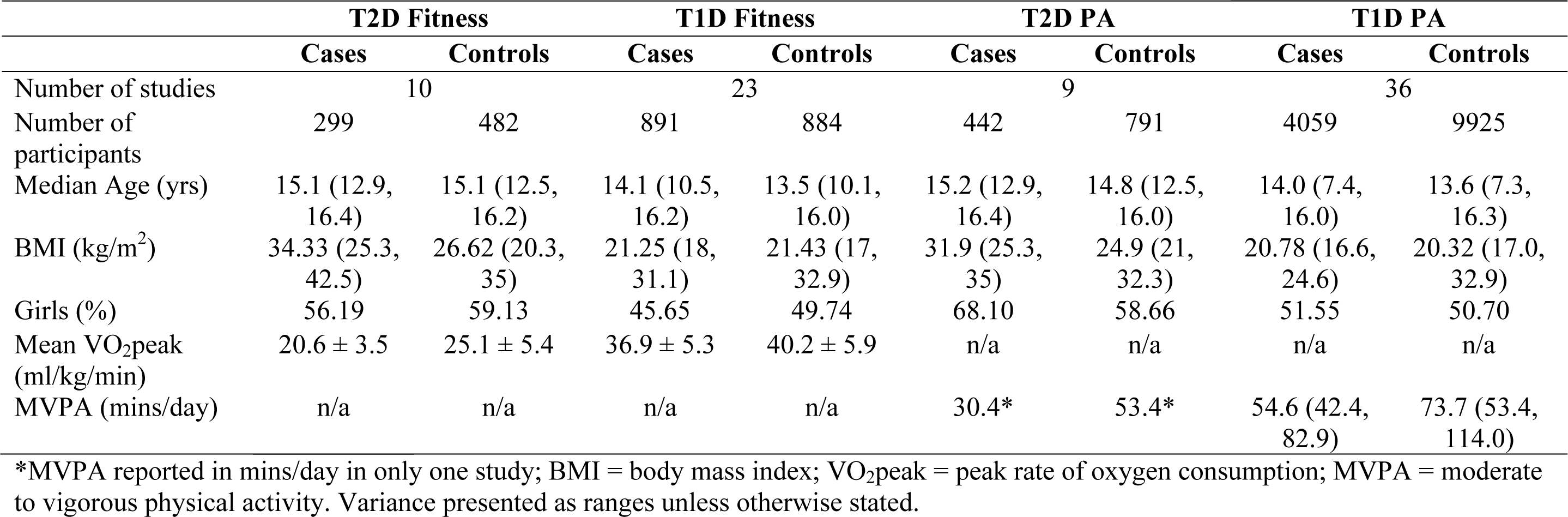
Details for the studies included in the systematic review and meta-analysis.

**Table 2.** Risk of score summary scores across different populations and outcomes.

| Variable | Type 1 Diabetes |  | Type 2 Diabetes |  |
| --- | --- | --- | --- | --- |
|  | PA (n= 36) | CRF (n= 23) | PA (n= 9) | CRF (n= 10) |
| Objective/Direct | 41.67% | 91.30% | 44.44% | 90.00% |
| Sample Recruitment | 88.89% | 82.61% | 88.89% | 80.00% |
| Time of Year | 36.11% | 4.35% | 22.22% | 0.00% |
| Place of Recruitment | 75% | 69.57% | 77.78% | 70.00% |
| Sample Description (mean /3) | 2.86 | 2.78 | 2.89 | 2.90 |
| Attrition (mean /2) | 1.46 | 1.39 | 1.44 | 1.5 |
| Data Collection & Reduction (mean /5) | 2.59 | 3.78 | 1.44 | 3.20 |
| PA intensity /Max criteria definition | 61.11% | 69.57% | 33.33% | 50.00% |
| Results (# actually analyzed) | 100% | 100% | 88.89% | 100% |
| Control Recruitment Described | 77.78% | 69.57% | 88.89% | 70.00% |
| Eligibility Criteria Described | 66.67% | 86.96% | 55.56% | 80.00% |
| Address Bias | 38.89% | 39.13% | 33.33% | 20.00% |
| Report Confounding | 63.89% | 73.91% | 88.89% | 80.00% |
| Adjust for Confounding | 13.89% | 17.39% | 33.33% | 40.00% |
| Sex Disaggregated Data | 22.22% | 21.74% | 0.00% | 10.00% |
| Ethnicity Data (mean /3) | 0.43 | 0.17 | 1.33 | 0.70 |
| Total Score (mean /25) | 14.05 | 15.39 | 13.67 | 15.20 |

PA = physical activity; CRF = cardiorespiratory fitness.

## DISCUSSION

The main finding from this systematic review and meta-analysis was that youth with type 2 diabetes have very low cardiorespiratory fitness, with values ∼25-50% lower than age-matched controls without diabetes and modestly lower daily physical activity. The mean VO_2_peak for youth with type 2 diabetes, ∼20 ml/kg/min, is well below normative values for adolescents and established thresholds for optimal cardiometabolic risk profiles. Among youth with type 1 diabetes, both cardiorespiratory fitness and physical activity are modestly lower compared to age-matched peers without diabetes. The differences in physical activity between adolescents with diabetes and controls, are more evident when objective measures are used to quantify physical activity. Lastly, these estimates are at risk of bias due to significant limitations in study designs and reporting, that likely contributes to the high heterogeneity of effect sizes across studies.

Low cardiorespiratory fitness is associated with increased cardiometabolic risk factor clustering in adolescence and cardiovascular disease-related morbidity in adulthood^2,10,78,79^. The average VO_2_peak derived from direct lab-based measures for youth with type 2 diabetes was 20.6 ml/kg/min, a value ∼50% lower than healthy weight controls without diabetes. In contrast, adolescents with type 1 diabetes displayed a mean VO_2peak_ value of 36.9 ml/kg/min and a value ∼10% lower than controls without diabetes. These levels of cardiorespiratory fitness for youth with type 2 diabetes are well below mean values for representative samples of adolescents in the US (46.4 ± 0.4 ml/kg/min for boys; 38.7 ± 0.3 ml/kg/min for girls)^80^; Canada (50.1 ml/kg/min, 95%CI: 48.1-52.0 ml/kg/min for boys; 42.0 ml/kg/min, 95% CI: 41.2-42.7 ml/kg/min for girls)^81^ and Europe^82^, and below the minimal threshold at which the odds of cardiometabolic risk factor clustering increase 2-4 fold (<41.6 ml/kg/min for boys; <34.6 ml/kg/min for girls)^2^. These deficits in cardiorespiratory fitness in youth living with diabetes may therefore contribute partially to the cardiovascular disease risk factor clustering reported in adolescence^28^ and excessive cardiovascular disease-related morbidity observed in young adulthood^83^. These data also support clinical practice guidelines and expert opinions that cardiorespiratory fitness is impaired in youth with both type 1 and type 2 diabetes.

Engaging in daily physical activity is a cornerstone in the management of pediatric diabetes^11–15^. Despite the widely recognized benefits of physical activity, very few systematic reviews with meta-analyses have been published describing daily physical activity levels in youth living with diabetes. One meta-analysis of cross-sectional studies of adolescents with chronic diseases found no difference in moderate to vigorous physical activity among youth with type 1 diabetes, relative to a comparison population^17^. With an additional 5 studies of adolescents with type 1 diabetes that wore a device to quantify physical activity objectively, we found a very small difference in daily physical activity among youth with type 1 diabetes, relative to peers without diabetes. Fewer studies have documented levels of daily physical activity among youth with type 2 diabetes, however they consistently reveal very low levels of daily physical activity, relative to expert recommendations (∼7000 steps/day and ∼35 minutes of moderate to vigorous physical activity daily)^17,84–86^. Compared to youth without diabetes, the differences in daily physical activity were greater for youth with type 2 diabetes, compared to youth with type 1 diabetes. The large deficits in daily physical activity among youth with type 2 diabetes may contribute to the lower cardiorespiratory fitness reported here, reinforcing the need for novel approaches to support physical activity behaviours for adolescents with type 2 diabetes.

An important observation from the current systematic review was the significant risk of bias observed in most studies published to date. Specifically, few studies adequately report sampling strategies for recruiting cases and controls, they also consistently fail to use objective methods to quantify daily physical activity and rarely adhere to reporting guidelines when they are used. Additionally, despite calls for reporting sex- and ethnicity specific outcomes, there are limited data describing sex-specific or ethnicity specific differences in daily physical activity or cardiorespiratory fitness for youth living with diabetes. Well designed, properly controlled, descriptive epidemiological studies that describe differences in physical activity and cardiorespiratory fitness among for boys and girls living with diabetes, and across various racial and ethnic groups are needed to overcome these limitations.

### Limitations

The current systematic review is the largest and most comprehensive description of cardiorespiratory fitness and physical activity levels among youth living with diabetes to date. Although we applied strict criteria for study inclusion and published the methods a-priori, there are several limitations of the review to consider. First, the quality of a meta-analysis is dependent on the quality of studies included. As previously mentioned, the risk of bias was high in most studies, therefore the estimates provided here should be interpreted with caution. The external validity or transferability of the results is also limited by several factors, including failure to report race and ethnicity, socio-economic status and in most cases sex-specific differences in cardiorespiratory fitness and physical activity within each population. Structural factors that influence access, safety and participation in physical activity are important when studying differences in physical activity for Indigenous youth, youth of colour, new immigrants, girls and non-binary youth. Type 2 diabetes is more common among Indigenous, Black and Hispanic youth^24–26^ and those living in low income households^27^, however we were unable to describe disparities in cardiorespiratory fitness or physical activity across ethnicity, socio-economic position, sex or gender due to a lack of information for these demographic variables in the majority of studies published to date. Lastly, we did not include non-English publications or studies in the grey literature to increase feasibility of the review and this may have introduced selective reporting bias (e.g. publication bias).

In conclusion, youth living with type 2 diabetes display profoundly lower cardiorespiratory fitness and modestly lower daily physical activity compared to peers without diabetes. Deficits in cardiorespiratory fitness and daily physical activity are also evident, but less pronounced, among youth with type 1 diabetes. Studies of cardiorespiratory fitness and physical activity and among youth with diabetes suffer from significant design issues that increase the risk that these estimates are biased. Deficits in physical activity and cardiorespiratory fitness may contribute to the increased burden of cardiovascular disease morbidity in young adulthood in individuals diagnosed with diabetes in childhood or adolescents. These findings reinforce calls for novel interventions to empower youth living with diabetes to engage in regular physical activity and improve cardiorespiratory fitness.

## Data Availability

Data will be made available upon request.

## Abbreviations

T2D: type 2 diabetes
T1D: type 1 diabetes
CRF: cardiorespiratory fitness
PA: physical activity

## Acknowledgments

Drs McGavock, Rabbani and Abou-Setta had full access to all the data in the study and take responsibility for the integrity of the data and the accuracy of the data analysis.

